# Yemeni university students public perceptions toward the use of artificial intelligence in healthcare: A cross-sectional study

**DOI:** 10.1101/2024.02.27.24303457

**Authors:** Najmaddin A. H. Hatem, Mohamed Izham Mohamed Ibrahim, Seena A. Yousuf

## Abstract

The integration of artificial intelligence (AI) in healthcare has emerged as a transformative force, promising to enhance medical diagnosis, treatment, and overall healthcare delivery. Hence, this study investigates the university students perceptions toward using AI in healthcare. A cross-sectional survey was conducted at two major universities using a paper-based questionnaire from September 2023 to November 2023. Participants’ views regarding using artificial intelligence in healthcare were investigated using 25 items distributed across five domains. The Mann-Whitney U test was applied for the comparison of variables. The response rate for the survey was 75%, with a sample size of 279. More than half of the participants (52%, n = 145) expressed their belief in AI’s potential to reduce treatment errors in the future. However, about (61.6%, n = 172) of participants fear the influence of AI that could prevent doctors from learning to make correct patient care judgments, and it was widely agreed (69%) that doctors should ultimately maintain final control over patient care. Participants with experience with AI, such as engaging with AI chatbots, significantly reported higher scores in both the “Benefits and Positivity Toward AI in Healthcare” and “Concerns and Fears” domains (*p* = 0.024) and (*p* = 0.026), respectively. The identified cautious optimism, concerns, and fears highlight the delicate balance required for successful AI integration. The findings emphasize the importance of addressing specific concerns, promoting positive experiences with AI, and establishing transparent communication channels. Insights from such research can guide the development of ethical frameworks, policies, and targeted interventions, fostering a harmonious integration of AI into the healthcare landscape in developing countries.

## Introduction

Artificial intelligence (AI) is anticipated to become the gold standard for health care because of its power to analyze and understand massive amounts of data and make more precise, intelligent judgments than humans [1,2,3]. Expert systems, machine learning, deep learning, and artificial neural networks (ANNs) are some of the several shapes that AI may take. Incorporating AI into many healthcare services has been made possible by improvements in accessible storage, quicker networks, and more powerful computers. AI is now mature enough to be used in image analysis, illness pattern prediction, and triage across various medical specialties, including radiology, gynecology, neurology, cardiology, pathology, and robotic surgery [4]. Healthcare AI integration offers a unique opportunity to improve healthcare services.

AI has emerged as a developing trend in the healthcare sector due to the expansion of medical data availability and the creation of algorithms [5, 6]. AI can increase medical diagnostic accuracy by using semantic analysis and picture recognition. Stanford University research from 2017 found that AI outperformed human doctors in detecting skin cancer by achieving more than 90% diagnostic accuracy [7]. With the aid of ever-sophisticated algorithms and sensitive tools, AI technology can offer outstanding therapeutic options and carry out specialized procedures in the context of medical decision-making and therapy. For instance, the American Children’s National Health System research team built the first robotic hand to autonomously handle soft tissue [8]. Researchers from the Las Vegas Department of Health have created an AI system that can aid in preventing foodborne diseases by applying machine learning to Twitter data [9]. Richer medical and health data offer prospects for more precise disease prevention and health monitoring in preventive healthcare. Deep-Mind researchers recently created a model to anticipate acute kidney damage 48 hours before using this method [10].

Regardless, AI is rapidly integrated into healthcare and will significantly impact services. For AI to reach its full potential in health care, it must be adopted by physicians, health professionals, patients, and other members of the public. Understanding the public acceptability of technology is crucial in this regard. This includes attitudes and beliefs toward collecting and using health-related data, which is required for AI systems to work [11,12]. Furthermore, while experts agree on what is and is not AI in health care [11], public opinion may be more varied [12]. Effective engagement and adoption of AI technology in health care necessitate understanding what non-experts believe about the situation.

Given the speed of technological developments worldwide, understanding Yemen’s perspectives on AI is essential. The exploration of public perceptions regarding the use of AI in healthcare holds particular significance. Yemen faces unique challenges in healthcare accessibility and resources, exacerbated by ongoing civil conflict and humanitarian crises [13,14]. Investigating the perceptions of individuals in this context provides a crucial lens through which to understand how a population coping with limited access to medical resources perceives the potential of AI to address healthcare challenges. This study contributes to local considerations. It enriches the broader global understanding of AI integration in healthcare, shedding light on the dynamics of perception and acceptance in regions grappling with adversity [12]. The findings from Yemen may offer insights into tailoring strategies for AI implementation that are sensitive to the specific needs and concerns of populations facing resource constraints and complex healthcare landscapes.

Hence, this paper aims to explore university students perceptions of AI in healthcare, evaluate the factors influencing public attitudes, and discuss the implications for the successful implementation of AI technologies. By analyzing public opinion, understanding concerns, and addressing ethical considerations, we can ensure that AI in healthcare is embraced as a valuable tool to improve patient outcomes, enhance healthcare delivery, and foster a positive and inclusive healthcare ecosystem.

## Methods

### Study setting, design, and sampling

We conducted a cross-sectional study among general students at Hodeidah and Sana’a universities using a paper-based questionnaire. The inclusion criteria are the capability to read and understand the information in the questionnaire and the willingness to participate. There were no explicit exclusion criteria. The Raosoft sample size calculator (http://www.raosoft.com/samplesize.html) was used to estimate the sample size with a margin of error of 5%, a confidence interval of 95%, and a 50% response rate. The estimated sample size was 377. Hence, we randomly distributed 400 questionnaires; 200 were distributed at Hodeidah University, and the other 200 were distributed at Sana’a University.

### Participants

University general students often have the potential to shape public opinion and contribute to societal discussions. Their views on using AI in healthcare can have a ripple effect on the broader population, making their insights particularly valuable since the sample size involved students in any discipline from two major universities in two different multicultural cities in the country. By including students from various cities and universities, we can capture regional variations in public perception toward AI use in healthcare. Other cities may have varying levels of exposure to AI technologies, cultural influences, and healthcare infrastructure, which can shape perceptions.

### Development of the questionnaire

The questionnaire was adopted from the previous validated quantitative study [15]. The questionnaire was introduced in both Arabic and English. The questionnaire included three parts. The socio-demographic information on students, age, gender, etc., was represented in the first part. In the second part, 25 items were divided into five different domains “*Benefits and Positivity, Concerns and Fears, Trust and Confidence in AI and Doctors, Patient Involvement and Data Sharing, and Professional Autonomy of Doctors*”; participants were asked to rate on a 5-point Likert scale of (5= Strongly agree, 1= Strongly disagree), and those items represent participants’ perceptions concerning AI use in healthcare. The questionnaire was tested for its face and content validity. Three independent faculty members evaluated the questionnaire to determine the relevance, clarity, conciseness, and ease of understanding of the items assessed by four students. Experts’ comments were considered in the final draft of the questionnaire.

### Consent and ethical approval

Verbal consent was obtained from all participants included in the study. They were informed about the proposal of the study, their rights as participants, and the confidentiality of their data. Their voluntary participation was acknowledged, and they were assured that their identities would remain anonymous throughout the study. The study design and procedure were approved by the Institutional Review Board at Aden University, Aden, Yemen, in compliance with the International Conference of Harmonization (ICH) Research Code (REC-164-2023).

### Data analysis

The data was coded and analyzed using the statistical package for the social sciences (SPSS) version 26 (Armonk, NY: IBM Corp.). The Kolmogorov-Smirnov test was used to test normality. Frequencies, percentages, median, and IQR were used to determine the respondent’s socio-demographic characteristics. Frequencies and percentages were used to determine the participant’s perception of AI use in healthcare. The Mann-Whitney U test was applied for the comparison of variables.

## Results

### General results

The study comprised a sample size of 279 participants with a response rate of 74%, with the majority falling within the age range of 18–24 years (82.8%, n=231), while a smaller proportion comprised individuals aged 25 years and older (17.2%, n=48). Regarding gender distribution, the participants were divided into (38.4%, n=107) males and (61.6%, n=172) females. Regarding permanent residency, the majority resided in urban areas (84.6%, n=236), whereas a smaller percentage lived in rural regions (15.4%, n=43). When asked about their educational background, (44.1%, n=123) of participants identified as medical or other health-related specialties students. Furthermore, when questioned about their experience with artificial intelligence AI chatbots, (30.5%, n=85) confirmed utilizing them, whereas the majority (69.5%, n=194) had not yet engaged with such technology (Table 1). When participants asked about their overall extant of positivity toward the use of AI in healthcare, about half of them were positive Fig 1. In table 1 participants who have previously used AI chatbots reported higher scores in terms of the overall perceptions on AI use in Healthcare (*p* = 0.023).

**Fig 1.**
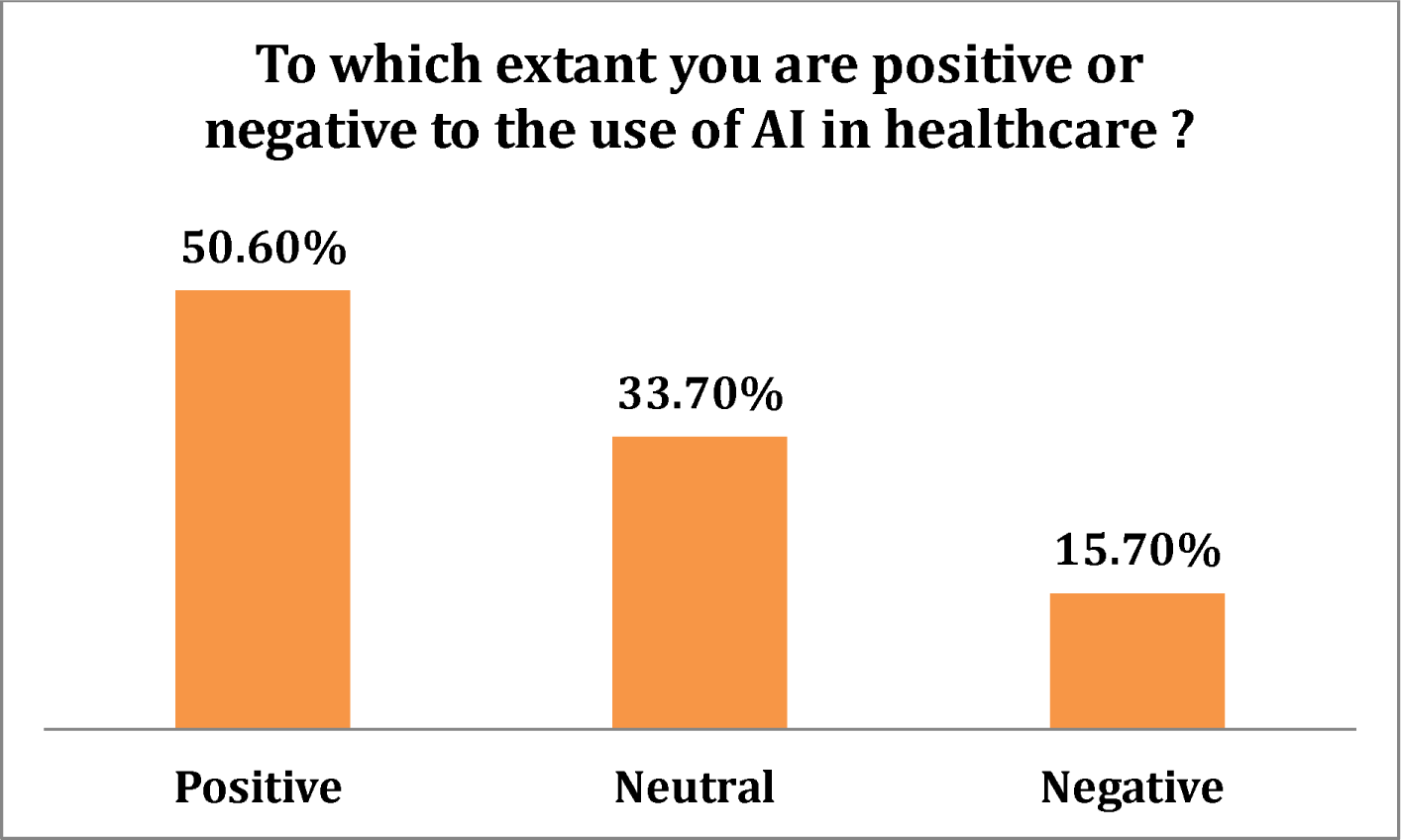
Participants positivity extant toward the use of AI in healthcare.

**Table 1.**
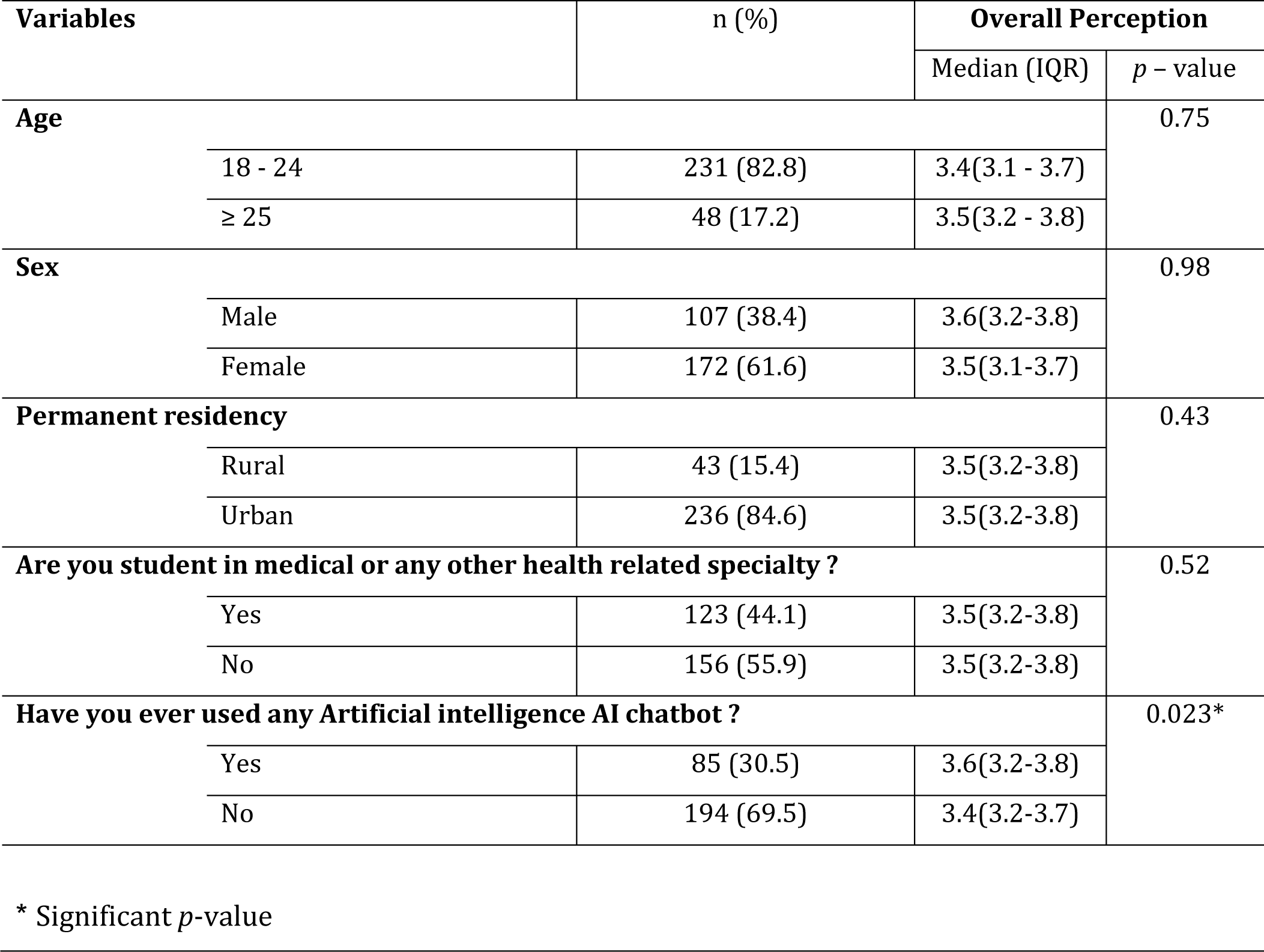
Participants’ Socio-demographics & characteristics.

### 1. Benefits and positivity toward AI in healthcare

Table 2 below shows moderate perceptions regarding *Benefits and Positivity Toward AI in the Healthcare* domain with a median and IQR of 3.5(3.2–3.8). Furthermore, the overall score of this domain was significantly affected by the use of AI chatbots, in which participants who have previously used AI chatbots reported higher scores with a median and IQR of 3.7(3.3–4) and (*p* = 0.024). (Table 3.)

As shown in Table 2, participants generally agreed that using artificial intelligence benefits patients. Furthermore, over half of the participants expressed their belief in AI’s potential to reduce treatment errors in the future. There was an agreement (over 60%, n=172) that AI can provide doctors with more dedicated time to emphasize patient care. Additionally, a significant majority (over three-fifths) of participants regarded AI as an effective tool to address the overload and shortage of doctors.

**Table 2.**
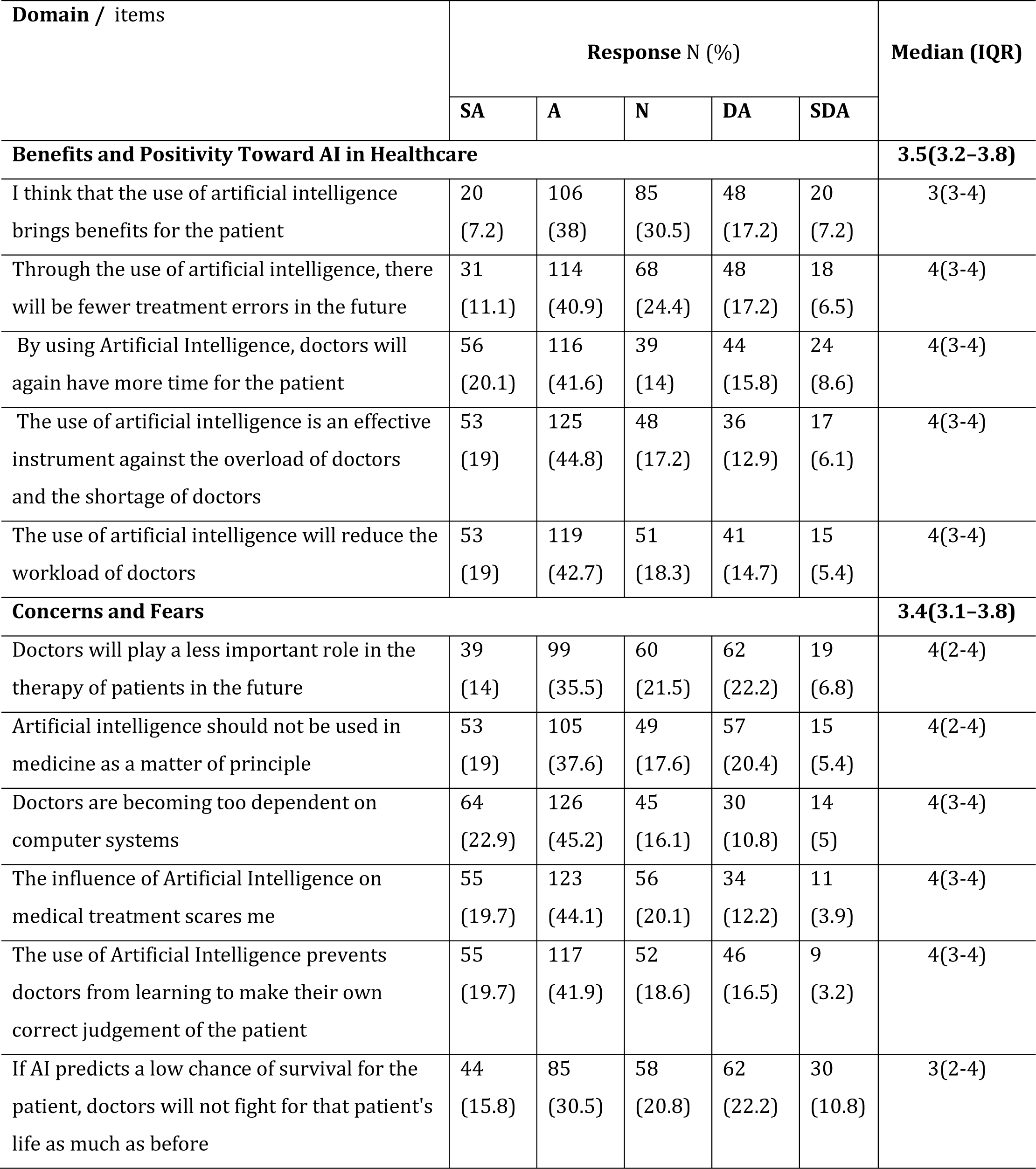
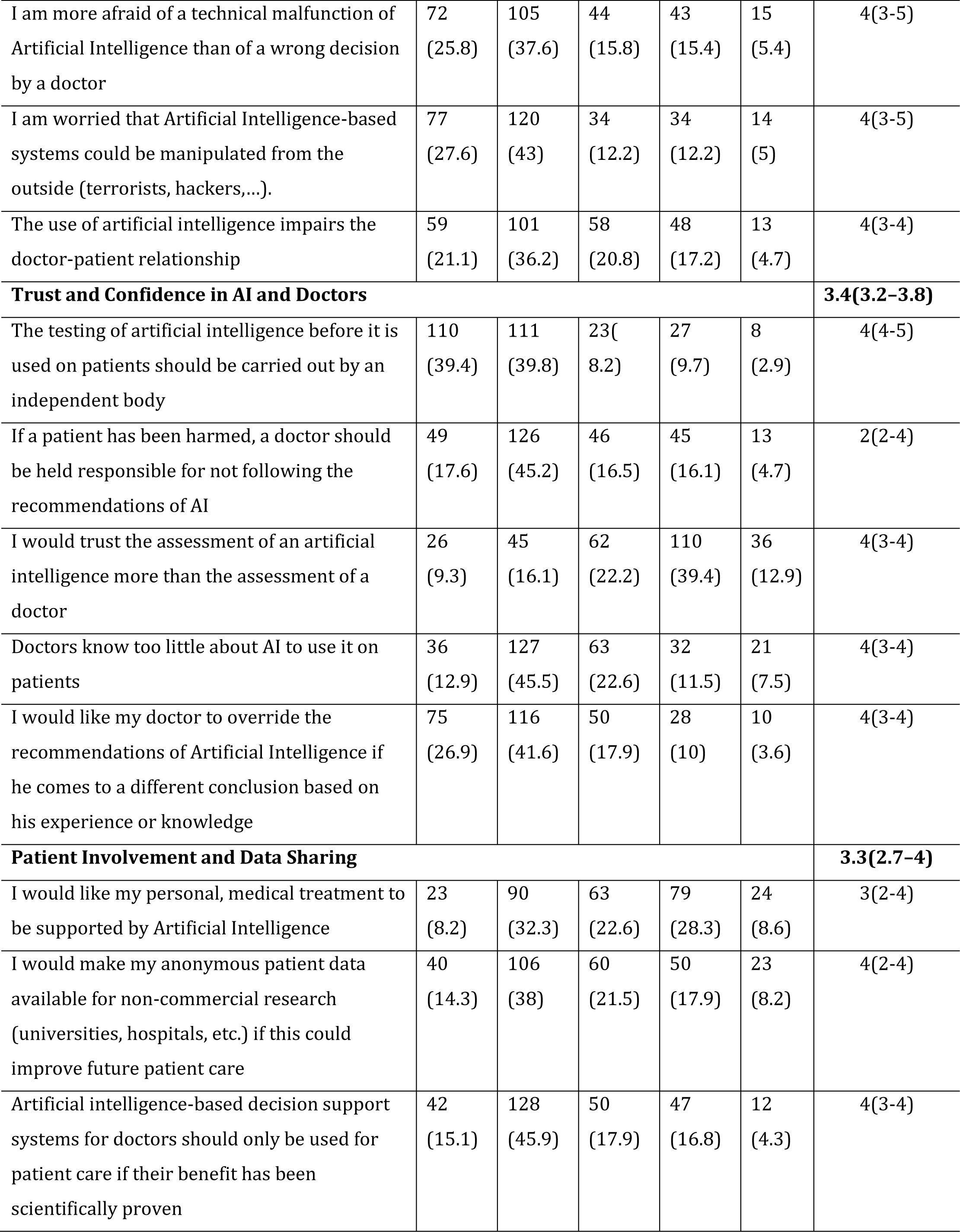
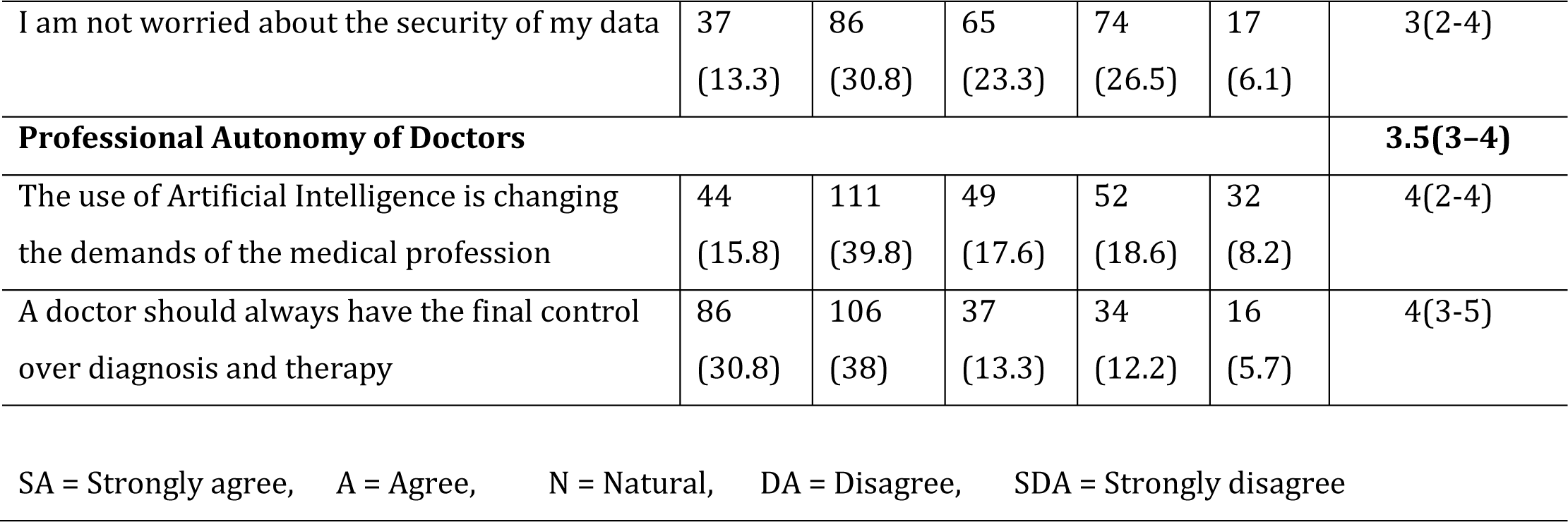
Participants’ perception of different domains of AI in healthcare.

| Domain / items | Response N (%) |  |  |  |  | Median (IQR) |
| --- | --- | --- | --- | --- | --- | --- |
|  | SA | A | N | DA | SDA |  |
| **Benefits and Positivity Toward AI in Healthcare** |  |  |  |  |  | **3.5(3.2–3.8)** |
| I think that the use of artificial intelligence brings benefits for the patient | 20<br>(7.2) | 106<br>(38) | 85<br>(30.5) | 48<br>(17.2) | 20<br>(7.2) | 3(3-4) |
| Through the use of artificial intelligence, there will be fewer treatment errors in the future | 31<br>(11.1) | 114<br>(40.9) | 68<br>(24.4) | 48<br>(17.2) | 18<br>(6.5) | 4(3-4) |
| By using Artificial Intelligence, doctors will again have more time for the patient | 56<br>(20.1) | 116<br>(41.6) | 39<br>(14) | 44<br>(15.8) | 24<br>(8.6) | 4(3-4) |
| The use of artificial intelligence is an effective instrument against the overload of doctors and the shortage of doctors | 53<br>(19) | 125<br>(44.8) | 48<br>(17.2) | 36<br>(12.9) | 17<br>(6.1) | 4(3-4) |
| The use of artificial intelligence will reduce the workload of doctors | 53<br>(19) | 119<br>(42.7) | 51<br>(18.3) | 41<br>(14.7) | 15<br>(5.4) | 4(3-4) |
| **Concerns and Fears** |  |  |  |  |  | **3.4(3.1–3.8)** |
| Doctors will play a less important role in the therapy of patients in the future | 39<br>(14) | 99<br>(35.5) | 60<br>(21.5) | 62<br>(22.2) | 19<br>(6.8) | 4(2-4) |
| Artificial intelligence should not be used in medicine as a matter of principle | 53<br>(19) | 105<br>(37.6) | 49<br>(17.6) | 57<br>(20.4) | 15<br>(5.4) | 4(2-4) |
| Doctors are becoming too dependent on computer systems | 64<br>(22.9) | 126<br>(45.2) | 45<br>(16.1) | 30<br>(10.8) | 14<br>(5) | 4(3-4) |
| The influence of Artificial Intelligence on medical treatment scares me | 55<br>(19.7) | 123<br>(44.1) | 56<br>(20.1) | 34<br>(12.2) | 11<br>(3.9) | 4(3-4) |
| The use of Artificial Intelligence prevents doctors from learning to make their own correct judgement of the patient | 55<br>(19.7) | 117<br>(41.9) | 52<br>(18.6) | 46<br>(16.5) | 9<br>(3.2) | 4(3-4) |
| If AI predicts a low chance of survival for the patient, doctors will not fight for that patient's life as much as before | 44<br>(15.8) | 85<br>(30.5) | 58<br>(20.8) | 62<br>(22.2) | 30<br>(10.8) | 3(2-4) |
| I am more afraid of a technical malfunction of Artificial Intelligence than of a wrong decision by a doctor | 72<br>(25.8) | 105<br>(37.6) | 44<br>(15.8) | 43<br>(15.4) | 15<br>(5.4) | 4(3-5) |
| I am worried that Artificial Intelligence-based systems could be manipulated from the outside (terrorists, hackers,...). | 77<br>(27.6) | 120<br>(43) | 34<br>(12.2) | 34<br>(12.2) | 14<br>(5) | 4(3-5) |
| The use of artificial intelligence impairs the doctor-patient relationship | 59<br>(21.1) | 101<br>(36.2) | 58<br>(20.8) | 48<br>(17.2) | 13<br>(4.7) | 4(3-4) |
| **Trust and Confidence in AI and Doctors** |  |  |  |  |  | **3.4(3.2-3.8)** |
| The testing of artificial intelligence before it is used on patients should be carried out by an independent body | 110<br>(39.4) | 111<br>(39.8) | 23(<br>8.2) | 27<br>(9.7) | 8<br>(2.9) | 4(4-5) |
| If a patient has been harmed, a doctor should be held responsible for not following the recommendations of AI | 49<br>(17.6) | 126<br>(45.2) | 46<br>(16.5) | 45<br>(16.1) | 13<br>(4.7) | 2(2-4) |
| I would trust the assessment of an artificial intelligence more than the assessment of a doctor | 26<br>(9.3) | 45<br>(16.1) | 62<br>(22.2) | 110<br>(39.4) | 36<br>(12.9) | 4(3-4) |
| Doctors know too little about AI to use it on patients | 36<br>(12.9) | 127<br>(45.5) | 63<br>(22.6) | 32<br>(11.5) | 21<br>(7.5) | 4(3-4) |
| I would like my doctor to override the recommendations of Artificial Intelligence if he comes to a different conclusion based on his experience or knowledge | 75<br>(26.9) | 116<br>(41.6) | 50<br>(17.9) | 28<br>(10) | 10<br>(3.6) | 4(3-4) |
| **Patient Involvement and Data Sharing** |  |  |  |  |  | **3.3(2.7-4)** |
| I would like my personal, medical treatment to be supported by Artificial Intelligence | 23<br>(8.2) | 90<br>(32.3) | 63<br>(22.6) | 79<br>(28.3) | 24<br>(8.6) | 3(2-4) |
| I would make my anonymous patient data available for non-commercial research (universities, hospitals, etc.) if this could improve future patient care | 40<br>(14.3) | 106<br>(38) | 60<br>(21.5) | 50<br>(17.9) | 23<br>(8.2) | 4(2-4) |
| Artificial intelligence-based decision support systems for doctors should only be used for patient care if their benefit has been scientifically proven | 42<br>(15.1) | 128<br>(45.9) | 50<br>(17.9) | 47<br>(16.8) | 12<br>(4.3) | 4(3-4) |
| I am not worried about the security of my data | 37<br>(13.3) | 86<br>(30.8) | 65<br>(23.3) | 74<br>(26.5) | 17<br>(6.1) | 3(2-4) |
| **Professional Autonomy of Doctors** |  |  |  |  |  | **3.5(3-4)** |
| The use of Artificial Intelligence is changing the demands of the medical profession | 44<br>(15.8) | 111<br>(39.8) | 49<br>(17.6) | 52<br>(18.6) | 32<br>(8.2) | 4(2-4) |
| A doctor should always have the final control over diagnosis and therapy | 86<br>(30.8) | 106<br>(38) | 37<br>(13.3) | 34<br>(12.2) | 16<br>(5.7) | 4(3-5) |

SA = Strongly agree, A = Agree, N = Natural, DA = Disagree, SDA = Strongly disagree

### 2. Concerns and fears

The *Concerns and Fears* domain in the current results showed a median and IQR of 3.4(3.1–3.8). As shown in Table 2, about half of the participants were concerned about the potential for doctors to play a less important role in patient therapy in the future, and over two-thirds of the participants expressed their belief that doctors are becoming excessively dependent on computer systems. Approximately (64%, n=178) of participants feared AI’s influence on medical treatment. Furthermore, concerns were raised by almost two-thirds of the participants regarding technical malfunctions and the potential manipulation of AI-based systems. Around (70%, n=197) of participants expressed worries worried about external entities such as terrorists and hackers manipulating AI-based systems. Moreover, a notable concern arose among some participants (46.3%, n=139), who feared doctors might be less motivated to fight for a patient’s life if AI predicts a low chance of survival. More than half of the participants believe that using AI could potentially undermine the doctor-patient relationship. In addition, more than three-fifths of respondents felt that AI limits doctors’ autonomy in rendering their judgments.

**Table 3.** Analysis of five domains of perception of AI in healthcare.

| Variables | Benefits and Positivity Toward AI in Healthcare |  | Concerns and Fears |  | Trust and Confidence in AI and Doctors |  | Patient Involvement and Data Sharing |  | Professional Autonomy of Doctors |  |
| --- | --- | --- | --- | --- | --- | --- | --- | --- | --- | --- |
|  | Median (IQR) | *P - Value* | Median (IQR) | *P - Value* | Median (IQR) | *P - Value* | Median (IQR) | *P - Value* | Median (IQR) | *P - Value* |
| Age |  |  |  |  |  |  |  |  |  |  |
| 18 – 24 | 3.5<br>(3.2-3.8) | 0.70 | 3.4<br>(3.1-3.8) | 0.07 | 3.4<br>(3.2-3.8) | 0.5 | 3.3<br>(2.6-4) | 0.5 | 3.5<br>(3-4) | 0.84 |
| ≥ 25 | 3.5<br>(3-4) |  | 3.6<br>(3.2-4) |  | 3.6<br>(3-4) |  | 3.3<br>(2.6-3.6) |  | 3.5<br>(3-4.2) |  |
| Sex |  |  |  |  |  |  |  |  |  |  |
| Male | 3.5<br>(3.2-4) | 0.39 | 3.4<br>(3.2-3.8) | 0.43 | 3.6<br>(3.3-4) | 0.023* | 3.3<br>(3-4) | 0.006* | 3.5<br>(3-4.5) | 0.52 |
| Female | 3.5<br>(3.2-3.8) |  | 3.4<br>(3.1-3.8) |  | 3.4<br>(3.3-3.8) |  | 3.3<br>(2.7-3.7) |  | 3.5<br>(3-4) |  |
| Permanent residency |  |  |  |  |  |  |  |  |  |  |
| Rural | 3.5<br>(3.2-4) | 0.67 | 3.4<br>(3.2-3.9) | 0.48 | 3.6<br>(3.2-3.9) | 0.25 | 3.3<br>(2.7-4) | 0.4 | 3.5<br>(3-4.3) | 0.81 |
| Urban | 3.5<br>(3.2-3.8) |  | 3.4<br>(3.1-3.7) |  | 3.4<br>(3.2- 3.8) |  | 3.3<br>(2.7-4) |  | 3.5<br>(3-4) |  |
| Are you student in medical or any other health related specialty ? |  |  |  |  |  |  |  |  |  |  |
| Yes | 3.5<br>(3.2-4) | 0.18 | 3.4<br>(3.2-3.8) | 0.24 | 3.4<br>(3 - 3.8) | 0.66 | 3.3<br>(2.7-4) | 0.56 | 3.5<br>(3-4.5) | 0.05 |
| No | 3.5<br>(3.2-3.8) |  | 3.4<br>(3-3.8) |  | 3.4<br>(3.2-3.9) |  | 3.3<br>(3-3.7) |  | 4<br>(3-4) |  |
| Have you ever used any Artificial intelligence AI chatbot ? |  |  |  |  |  |  |  |  |  |  |
| Yes | 3.7<br>(3.3-4) | 0.024* | 3.7<br>(3.2-3.9) | 0.026* | 3.6<br>(3.2-4) | 0.097 | 3.3<br>(2.7-4) | 0.62 | 4<br>(3-4.5) | 0.12 |
| No | 3.5<br>(3.2-3.8) |  | 3.4<br>(3.1-3.8) |  | 3.4<br>(3.2-3.8) |  | 3.3<br>(2.7-4) |  | 3.5<br>(3-4) |  |

\* Significant *p*-value

Table 3 indicated that the overall score of the *Concerns and Fears* domain was significantly affected by the use of AI chatbots, with participants who had previously used AI chatbots reporting higher scores (*p* = 0.026). In S1, participants studying health or medical specialties and those who previously used AI bots reported higher belief scores that doctors are excessively reliant on computer systems than their peers (*p* = 0.018, *p* = 0.032), respectively. Furthermore, AI chatbot users reported higher scores with a median and IQR of 4(4-5) to fear external entities such as terrorists and hackers manipulating AI-based systems (*p* = 0.046).

### 3. Trust and confidence in AI and doctors

A significant portion exceeding (60%, n=205), believed that if a patient experiences harm, the responsibility should lie with the doctor for not adhering to AI recommendations. Moreover, there existed a divergence in trust between AI assessments and doctors’ evaluations, with approximately (25%, n=71) of participants favoring AI assessments over those of doctors. In contrast, over (50%, n=146) contested this opinion, and 22.2% maintained a “neutral” stance. A notable percentage of participants (79.2%, n=221) strongly supported testing AI before its use on patients by independent bodies. Moreover, most participants (58.4%, n=163) expressed apprehension about doctors’ limited knowledge and understanding of AI’s application in patient care.

In S1, participants studying health or medical specialties reported higher belief scores regarding the trust of AI assessments over doctors’ evaluations, with median and IQR of 3(2–4) and 2(2–3) for participants in other non-health-related specialties, respectively (*p* = 0.036). AI chatbot users reported higher scores with a median and IQR of 5(4–5) to the idea of testing AI before its usage on patients by an independent body (*p* = 0.005).

### 4. Patient involvement and data sharing

This domain reported the lowest perception score compared to other domains reported in this study, with a median and IQR of 3.2(2.7–4). Table 2 revealed that a considerable number (40.5%, n=113) of participants expressed a desire for their medical treatment to be supported by AI. Furthermore, it was important for many participants (61%, n=170) that AI-based decision support systems should have scientifically proven benefits for patient care. Interestingly, (44.1%, n=123) of participants expressed no worries about their data security.

### 5. The professional autonomy of doctors

Our data results are shown in Table 2. It is reported that about (55.6%, n=155) of respondents recognized that AI is significantly altering the demands of the medical profession. However, it was widely agreed (69%, n=192) that doctors should ultimately maintain final control over patient care.

## Discussion

The study aimed to explore university students perceptions regarding using artificial intelligence (AI) in healthcare. The findings across different domains - *Benefits and Positivity, Concerns and Fears, Trust and Confidence in AI and Doctors, Patient Involvement and Data Sharing, and Professional Autonomy of Doctors* - provide valuable insights into the complex landscape of perceptions toward AI in healthcare. It’s essential to acknowledge the context in which these perceptions were obtained, given the challenges of healthcare accessibility and resources in Yemen. Yemen faces significant healthcare challenges, including limited access to medical resources and facilities [13,14]. As a result, participants may be inclined to view AI as a potential solution to improve healthcare delivery and outcomes in the face of these challenges.

Our findings indicate a cautious optimism among participants regarding the benefits and positivity of AI in healthcare. Despite concerns and fears, such as the potential diminishing role of doctors and worries about technical malfunctions, the study highlights the participants’ acknowledgment of AI’s potential to address healthcare challenges in Yemen, where there are limited resources and ongoing conflicts. Notably, the study emphasizes the impact of experience with AI applications, particularly AI chatbots, on shaping perceptions, suggesting the importance of exposure and education. The results also reveal a complex relationship between trust in AI and doctors, with a significant portion of participants holding doctors responsible for harm from not adhering to AI recommendations.

### Benefits and positivity toward AI in healthcare

Our study indicated that participants had modest perspectives on the advantages and positives of AI in healthcare. This implies a cautious optimism among individuals, implying that while they see the potential benefits of AI in healthcare, they may still have doubts or misgivings [12]. This study emphasizes the need for more research and education on AI in healthcare to overcome any misconceptions or apprehensions that the public may have. However, participants in our survey largely agreed that AI might assist patients by lowering treatment mistakes and doctors’ workload and giving doctors more time to emphasize patient care. This is similar to recent findings from Nigerian and German studies [16,15]. This acknowledgement of AI’s potential beneficial influence on healthcare delivery demonstrates that they perceive the worth of implementing AI technology. This optimistic mindset might originate from a desire to close current gaps in the healthcare system and improve patient safety. It might also result from increased faith in technology to solve complex healthcare challenges [12,17,11]. Furthermore, the participants’ perspectives reflect a practical awareness of Yemen’s healthcare situation, with AI identified as a viable option to address the issues of inadequate healthcare resources and a physician shortage. Furthermore, our survey found that participants believe AI can decrease treatment mistakes and provide more committed time for patient care. This aligns with the global aim of improving healthcare outcomes through technological advances [4].

Another interesting conclusion from the study is that AI chatbot usage considerably impacts overall perception scores. This suggests that experience with AI applications, such as AI chatbots, might lead to more positive impressions of AI in healthcare. This conclusion highlights the need to allow people to interact with AI systems in hospital settings to create familiarity, knowledge, and acceptance.

### Concerns and fears

About half of the participants expressed concern about the possibility of doctors playing a lesser role in patient therapy in the future, highlighting concerns about the changing dynamics in healthcare delivery. This worry reflects a realization of AI’s transformational potential in healthcare and a desire to retain the human touch in medical treatment as technology advances. Furthermore, this issue may originate from the idea that artificial intelligence systems may completely replace human doctors. A striking example of this issue is the study conducted in China among users of “Sina Weibo,” a Chinese social media platform using mixed methods. Among 200 (21%) reported posts mentioning AI replacing human doctors, 95 (47·5%) expressed that it would do so completely [18]. While AI can help with diagnostic and treatment decisions, it is critical to understand that physicians provide a depth of knowledge, empathy, and ethical judgment that AI cannot replace.

The results of the current study showed that close to two-thirds were concerned about using artificial intelligence (AI) in medical treatment. This apprehension derives from restricted access to healthcare facilities and resources in Yemen, a weaker healthcare system as a result of the ongoing civil conflict and humanitarian catastrophe, and fears about the possible loss of human touch and customized treatment. Furthermore, a lack of understanding regarding AI technology and its limits may add to misconceptions and concerns, particularly when entrusting AI with vital medical decisions. Moreover, existing societal and cultural views in Yemen may add to the fear. Some viewed AI’s usage as interfering with the natural order or performing a function reserved for human professionals. These worries originate from potential ethical issues and a concern about not completely controlling or comprehending AI system judgments.

The result that approximately half of the participants in the study expressed fear regarding doctors being less motivated to fight for a patient’s life if AI predicts a low chance of survival raises significant concerns about the ethical implications of using AI in healthcare; their proportion was much similar to a previous study conducted among German patients [15]. Firstly, this fear highlights the potential loss of trust between patients and healthcare professionals. Patients rely on their doctors to provide the best care and prioritize their well-being. Suppose AI is seen as influencing doctors’ decisions and potentially leading to a decline in their motivation to save a patient’s life. In that case, it can erode patient trust and negatively impact doctor-patient relationships. Secondly, this result raises questions about how AI predictions should be integrated into medical decision-making processes. While AI has the potential to enhance patient care and improve outcomes by providing accurate predictions, it should not replace the human judgment and empathy that physicians bring to their practice. Doctors have a moral and professional obligation to act in the best interest of their patients, irrespective of AI predictions.

Furthermore, the study reveals that approximately 63% of respondents were more afraid of a technical malfunction of AI than of a wrong decision by a doctor. This fear could be attributed to technology’s perceived unpredictability and potential dangers. The participants may have concerns about the reliability and accuracy of AI systems, worrying that a malfunction or an error in the algorithm could have severe consequences for patient outcomes. A common finding in many studies is that patients want physician supervision of AI and prefer a physician in a direct comparison [15,12,16,19]. A particularly striking example of the high respect and trust in the competence of physicians compared to AI tools is provided by the study of York and colleagues using the example of radiographic fracture identification. 24 On a 10-point scale representing confidence, the study population of 216 respondents awarded their human radiologists a near maximum score of 9.2 points, while the AI tool received only 7.0 points [20]. These findings suggest a need for transparency and clear communication about the limitations and capabilities of AI in healthcare. It is crucial to ensure that patients and healthcare professionals are well-informed about the functioning of AI systems and the safeguards to mitigate potential risks.

In addition, our study highlighted that participants with experience with AI applications, such as engaging with AI chatbots, might have more concerns and fears regarding the use of AI in healthcare.

### Trust and confidence in AI and doctors

The results indicate that a significant proportion of Yemeni participants hold doctors responsible for any harm experienced by patients due to their failure to adhere to AI recommendations, which raises critical concerns within the specific socio-cultural and healthcare context of the region. In Yemen, where healthcare resources and infrastructure may face challenges, the reliance on doctors is already pronounced. Hence, participants may be inclined to view AI as a potential solution to improve healthcare delivery and outcomes in the face of these challenges. Moreover, this suggests the high reliance on AI systems in healthcare decision-making is due to their potential to provide accurate and reliable recommendations. However, medical decision-making involves factors like patient history, personal preferences, clinical expertise, and contextual nuances [4,17]. Doctors are trained to exercise clinical judgment and integrate AI technologies with their knowledge and skills. While AI technologies can provide valuable insights, they should not be viewed as infallible or all-encompassing [17]. Doctors must balance AI recommendations with patient-specific considerations and their clinical judgment.

The finding that only 25% of participants preferred AI’s evaluation above that of physicians indicates a rising degree of user confidence and trust in AI technology. This may be explained by the potential advantages that AI may provide, such as its speedy analysis of enormous volumes of data and capacity to make suggestions based on patterns and trends. This participant group could see AI as a trustworthy and objective information source. However, it’s interesting to note that more than half of the participants disagreed with this statement, expressing doubt about using AI alone to make medical judgments. This mistrust may stem from worries about the drawbacks and hazards of AI technology, such as its inability to simulate human empathy and the possibility of biases or mistakes in its algorithms. These participants could be drawn to the particular knowledge, wisdom, and intuition that human physicians offer in decision-making. These findings are in line with extensively discussed reported results from different settings in a mixed methods systematic review that the overall participants trusted AI less than humans; however, this gap in trust was reduced when participants were able to choose between AI and humans freely [12].

The continuous discussion in medical care about how to strike the right balance between human judgment and AI suggestions is reflected in the differences in trust between AI and doctors’ assessments. It emphasizes the importance of properly weighing the benefits and drawbacks of using AI and human doctors to make well-informed, patient-centered judgments [1,3,6,17]. Achieving ideal patient results and preserving patient confidence in the healthcare system requires carefully balancing AI technology and human skills.

### Patient involvement and data sharing

Our study found that the domain of *Patient Involvement and Data Sharing* received the lowest perception score, suggesting concerns about patient involvement in decision-making and data sharing. About 40.5% of participants desired AI-supported medical treatment, indicating trust in AI’s potential benefits. 61% of participants emphasized the need for evidence-based practice and a cautious approach to AI integration. 44.1% of participants expressed no concerns about data security, indicating trust in healthcare organizations. The reasons behind the low perception score could include concerns about losing control over treatment decisions, fear of data misuse, and skepticism about patient input. To improve patient involvement and data sharing, transparent communication channels should be established, including educating patients about AI’s benefits and limitations, ensuring informed consent, and implementing robust security measures. Patients should also be involved in their healthcare journey, fostering shared decision-making processes and promoting patient empowerment. Collaborative efforts between healthcare providers, AI developers, and policymakers can help establish guidelines and ethical frameworks for patient involvement and data sharing [15,16,18].

### The professional autonomy of doctors

The finding that more than half of respondents recognized the changing demands of the medical profession due to artificial intelligence can be justified by the increasing integration of AI technologies in healthcare. As AI applications become more prevalent, healthcare professionals and the general public will likely become more aware of the transformative effects on medical practices [1-6,17,19]. This awareness is particularly significant considering the current global trend towards integrating AI into various aspects of healthcare [12]. This perception aligns with the growing impact of AI in healthcare, prompting individuals to acknowledge and adapt to the evolving landscape of the medical profession [12,15,16,19].

The majority of respondents believe that doctors should have the final say in diagnosis and therapy, despite AI systems, raising questions about the role of AI in healthcare. This study’s results are not completely surprising as they fit with most past research on patient and general public opinions; human physicians are typically trusted and followed more than AI systems [15,16,18,19,12]. This may be due to the perception that doctors have a level of expertise and experience that AI systems cannot replicate. AI systems are often seen as tools to support doctors in their decision-making process, but they do not replace them entirely. There are also concerns about the reliability and accuracy of AI systems, as they can produce incorrect or biased results. The complexity of diagnosis and therapy, involving various symptoms, patient history, and clinical data, may also influence this perception [1,3,4]. However, AI systems can rapidly analyze vast amounts of medical data, identify patterns, and generate insights, assisting doctors in making more informed decisions and providing timely, personalized care. AI tools can also improve healthcare access and efficiency [1-8,17]. The optimal approach is to balance human expertise and AI capabilities, incorporating AI insights into doctors’ decision-making while maintaining responsibility for the final diagnosis and therapy.

### Study limitations and strengths

While our study provides valuable insights into public perceptions of AI in healthcare among university students in Yemen, it is essential to acknowledge certain limitations. The sample primarily consists of university students, limiting the generalizability of the findings to a broader population. Additionally, the study’s cross-sectional nature captures a snapshot in time, and perceptions may evolve over the long term. The study’s strengths lie in its comprehensive exploration of diverse domains and the nuanced understanding it offers. Despite limitations, these findings serve as a foundation for targeted interventions and highlight the need for ongoing research to capture the dynamic nature of attitudes toward AI in Yemen’s healthcare landscape and other settings with similar contexts and resources.

### Study implications

The study on public perceptions of AI use in healthcare among Yemeni university students provides crucial insights that carry profound implications for the future of healthcare in the country. The identified cautious optimism, concerns, and fears emphasize the need for nuanced strategies for integrating AI technologies. To ensure successful implementation, initiatives should address specific concerns, promote positive experiences with AI, and establish transparent communication channels. Furthermore, the study underscores the importance of ongoing research, including longitudinal and cross-sectional studies, to track changing perceptions and explore variations across demographics. Insights from such research can guide the development of ethical frameworks, policies, and targeted interventions, fostering a harmonious integration of AI into the healthcare landscape in Yemen.

### Study recommendations

Below are a few recommendations in relation to the study findings:

- Increase participants’ awareness and understanding of AI in healthcare through educational programs, workshops, and information campaigns.
- Encourage further research and development in AI applications in healthcare that are tailored to the specific context and challenges faced in Yemen.
- Provide opportunities for individuals to interact with AI systems in hospital settings, such as AI chatbots, to create familiarity, knowledge, and acceptance. Positive experiences with AI may contribute to more favorable perceptions.
- Develop guidelines for the ethical integration of AI in medical decision-making processes to ensure that AI complements, rather than replaces, the judgment and empathy of human doctors.
- Address concerns about potential conflicts of interest between AI predictions and doctors’ motivations, emphasizing the importance of maintaining trust in doctor-patient relationships.
- Enhance transparency in AI technology by providing clear information about its limitations, potential risks, and the safeguards in place to mitigate those risks.
- Foster open communication between healthcare providers, AI developers, policymakers, and the public to address concerns and build trust.
- Establish transparent communication channels to educate patients about the benefits and limitations of AI, ensure informed consent, and address concerns about data security.
- Promote collaborative efforts between healthcare providers, AI developers, and policymakers to establish ethical frameworks for patient involvement and data sharing.
- Emphasize the complementary role of AI as a tool to support doctors in their decision-making process rather than replace them entirely.
- Encourage ongoing dialogue about the evolving role of AI in healthcare, highlighting the strengths of both AI systems and human doctors in providing optimal patient care.
- Recognize that perceptions of AI in healthcare may evolve, and therefore, continuous monitoring and research are essential to capture changing attitudes.
- Consider expanding the study to include a more diverse sample beyond university students to gain a broader understanding of public perceptions.
- Foster international collaborations to share best practices and experiences in integrating AI into healthcare, especially in regions facing similar challenges as Yemen.
- Advocate for the development of policies that ensure responsible and ethical use of AI in healthcare, taking into account the unique challenges and contexts of Yemen’s healthcare system.
- Plan for long-term interventions that address the evolving landscape of attitudes toward AI in healthcare, considering the dynamic nature of technology and public perceptions.

## Conclusion

Our study sheds light on the complex landscape of university students perceptions regarding the use of artificial intelligence (AI) in healthcare among participants in Yemen. The findings indicate a cautious optimism among participants regarding the benefits and positivity of AI in healthcare; however, this gap in trust was reduced when participants were able to freely choose between AI and humans. Despite concerns and fears, such as the potential diminishing role of doctors and worries about technical malfunctions, the study highlights the participants’ acknowledgment of AI’s potential to address healthcare challenges in Yemen, where there are limited resources and ongoing conflicts. Notably, the study emphasizes the impact of experience with AI applications, particularly AI chatbots, on shaping perceptions, suggesting the importance of exposure and education. The results also reveal a complex relationship between trust in AI and doctors, with a significant portion of participants holding doctors responsible for harm resulting from not adhering to AI recommendations. The lowest perception score in the *Patient Involvement and Data Sharing* domain underscores concerns about control over treatment decisions and data security. Insights from such research can guide the development of ethical frameworks, policies, and targeted interventions, fostering a harmonious integration of AI into the healthcare landscape in many developing countries.

## Data availability

The data that support the findings of this study are available from the first author, upon reasonable request.

## Author’s contributions

**Hatem NAH.** Conceptualization; Formal analysis; Methodology; Project administration; Resources; Writing – original draft and Writing – review & editing. **Mohamed Ibrahim MI.** Supervision and Writing – review & editing. **Yousuf SA.** Methodology; Resources and Writing – review & editing.

## Data Availability

All data produced in the present study are available upon reasonable request to the authors.

